# HIV testing and prevalence in fishing communities in rural Uganda: a cross-sectional study of 3197 individuals within SchistoTrack

**DOI:** 10.1101/2025.07.24.25332115

**Authors:** Hanh Lan Bui, Lauren Wilburn, Salim W. Nsimbe, Betty Nabatte, Geoffrey W. Oromcan, Raymond Mujuni, Juma Nabonge, Narcis B. Kabatereine, Adrian D. Smith, Goylette F. Chami

**Affiliations:** Big Data Institute, Nuffield Department of Population Health, University of Oxford, Oxford, United Kingdom; Uganda Institute of Allied Health and Management Sciences, Kampala, Uganda; Division of Vector Borne Diseases and Neglected Tropical Diseases, Uganda Ministry of Health, Kampala, Uganda; Pakwach District Local Government, Pakwach, Uganda; Buliisa District Local Government, Buliisa, Uganda; Mayuge District Local Government, Mayuge, Uganda; Nuffield Department of Population Health, University of Oxford, Oxford, United Kingdom

**Keywords:** HIV, Uganda, mobile, surveillance, care cascade, fishermen, fisherfolk, fishing, community

## Abstract

**Introduction:** Fisherfolk are recognised as a priority population for HIV control in generalised epidemic settings, yet information on differences in testing, diagnosis, and care cascade engagement compared to the general population remain limited.

**Methods:** We conducted a cross-sectional analysis of 3197 participants aged 5-92 years within the SchistoTrack community-based cohort in rural Uganda. From 15 January to 12 February 2024, participants from 52 shoreline villages in Pakwach, Buliisa, and Mayuge districts were tested for HIV using rapid diagnostic tests following the national testing algorithm and completed a structured HIV history survey with a district HIV counsellor. Six definitions of fisherfolk were assessed among 1931 adult participants using univariate and adjusted logistic regressions (controlling for age, gender, and district) of ever testing for HIV, testing in the past 12 months, and HIV status. Backward stepwise selection based on the lowest Bayesian Information Criterion was used to select fisherfolk and other participant variables.

**Results:** Overall, 6.94% (134/1931) of adult participants were living with HIV (PLHIV), of whom 22.39% (30/134) were newly diagnosed. 6.02% (25/415) of adults reporting fishing activities were HIV-positive. Of those, 80% (20/25) were status-aware, 76% (19/25) were on ART, and 100% (8/8) of those who knew their viral load reported viral suppression. No significant differences in care cascade engagement were found between PLHIV reporting fishing activities and the general population. Measured viral suppression was 70.59% (72/102) among PLHIV, with no significant differences by fishing activities. Fishing activities were significantly associated with higher odds of ever testing for HIV (OR 1.76, 95% CI 1.22-2.54), but not with testing in the past 12 months or HIV status. No consistent district-level differences were observed.

**Conclusion:** Individuals reporting fishing activities had higher lifetime testing and comparable HIV prevalence and care cascade engagement to the general population, although neither population met UNAIDS 95-95-95 targets.

## INTRODUCTION

In 2023, sub-Saharan Africa (SSA) accounted for nearly 65% of the global human immunodeficiency virus (HIV) burden, with 39.9 million people living with HIV (PLHIV) worldwide [1]. The HIV epidemic in SSA is more generalised than in other regions of the world, although key populations, such as men who have sex with men, people who inject drugs, and sex workers, now account for 51% of new infections [2, 3]. The HIV response in SSA is guided by the UNAIDS 95-95-95 targets, which aim to end the AIDS epidemic by 2030 [4]. World Health Organization (WHO) policy advises that all individuals diagnosed with HIV should have immediate access to antiretroviral therapy (ART) [5]. Increasing status awareness among PLHIV through timely testing is the entry point to accessing the HIV care cascade [6].

Countries in SSA have identified priority populations in need of targeted efforts for HIV testing and linkage to care [7]. In many SSA countries, including Uganda, an important priority population are fisherfolk – residents of fishing communities – comprising fishermen, fishmongers, and others involved in the fishing trade [8]. HIV prevalence in fisherfolk is heterogeneous across SSA and is estimated at 15-40% in Uganda, primarily assessed in fishing communities around Lake Victoria [7-9]. HIV testing coverage among fisherfolk also varies widely across studies and contexts; Opio et al. reported 54% in 2013, while the Rakai Community Cohort Study recorded an increase from 68% in 2011-2012 to 96% in 2016-2017 following provision of HIV testing and counselling [8, 10, 11].

Fisherfolk in Uganda and elsewhere are defined as a priority population for HIV services due to multiple structural, behavioural and sociodemographic barriers to consistent engagement with care [8, 12-16]. High levels of mobility, driven by seasonal fishing cycles and movement between landing sites, are a key factor contributing to their heightened vulnerability to HIV [8, 12, 15]. Mobility is gendered: men typically migrate seasonally for fishing, often spending months moving between fishing villages, while women involved in fish trade tend to move less frequently [16]. Fishing communities are often located in remote, under-resourced areas, with long distances to health facilities, inadequate transportation, limited access to healthcare, and poor healthcare infrastructure [8, 13, 15]. These structural barriers impede access to HIV testing and care, while risk behaviours such as transactional and commercial sex and alcohol use add to HIV acquisition risk among fisherfolk [8, 14].

While community-based outreach initiatives, including HIV self-testing and mobile outreach, have sought to mitigate structural barriers, data to evidence their effectiveness remains limited [12, 17, 18]. There is therefore a need to establish care cascades among fisherfolk and to contrast these with general populations. Further, HIV interventions and research with fishing communities have generally been conducted at the community level, sampling from landing sites where the entire village or community is treated as one unit [10-12, 19, 20]. However, it is unclear whether such ecological associations with higher HIV prevalence or lower testing among fisherfolk also hold at the individual level, underscoring the need for more precise definitions to effectively target this priority group in HIV interventions. Differences in sampling across studies make it challenging to assess the true prevalence of testing and PLHIV among the priority population of fisherfolk [10-12, 19, 20].

Here we examined HIV prevalence, testing coverage, and the care cascade in fishing communities in rural Eastern and Western Uganda. A total of 3197 individuals aged 5-92 were screened for HIV using the Uganda Ministry of Health national testing algorithm within the community-based cohort, SchistoTrack. The overall aims of our study were to compare HIV testing coverage, prevalence, and care cascade engagement between fisherfolk and the general population, and to assess the relevance of individual- and community-level definitions of fisherfolk in understanding variation in HIV status and testing.

## METHODS

### Study context

A nested cross-sectional study was conducted within the community-based SchistoTrack cohort, which examined participants aged 5 years and older from 52 shoreline villages in Pakwach, Buliisa and Mayuge districts [21, 22]. HIV testing was introduced during the 2024 annual cohort follow-up following observation of a correlation between self-reported HIV status and periportal fibrosis [23]. From 15 January to 12 February 2024, study participants were tested for HIV using rapid diagnostic tests following the national algorithm and completed a structured HIV history survey with a district HIV counsellor. Viral load was subsequently collected for a subset of HIV-positive participants 12 months later.

### Outcomes

The primary outcomes of the study were lifetime HIV testing (ever testing) and current HIV infection status (both binary). To compare the determinants of testing recency with those of ever testing, we also examined HIV testing in the past 12 months among participants who self-reported being HIV-negative. HIV status was determined according to the Uganda Ministry of Health national testing algorithm based on three tests: Determine HIV-1/2 (Abbott) as a first-pass screening test, HIV 1/2 STAT-PAK Assay (Chembio Diagnostics) as a confirmatory test, and Bioline HIV 1/2 3.0 (Abbott) as a tie-breaker where the first two tests disagreed (Figure S1) [9].

The care cascade measures among PLHIV were self-reported in the 2024 HIV history survey and included HIV status awareness, linkage to care (defined as seeing a healthcare provider for the first time within 30 days of diagnosis), ART initiation (ever taking ART to treat HIV), current ART use, retention in care (defined as receiving HIV care from a healthcare provider in the past 12 months), and self-reported HIV viral load suppression (here defined as self-reported viral load of <1000 copies/mL) (Table S1). Additionally, PLHIV were asked to report the HIV clinic where they accessed ART; for newly diagnosed individuals, the clinic associated with their newly issued ART number was recorded. Participants who did not know or refused to report their lifetime testing history were excluded from all analyses (Figure 1). Those who did not know or refused to answer other self-reported outcomes were excluded only from analyses involving those specific outcomes.

**Figure 1.**
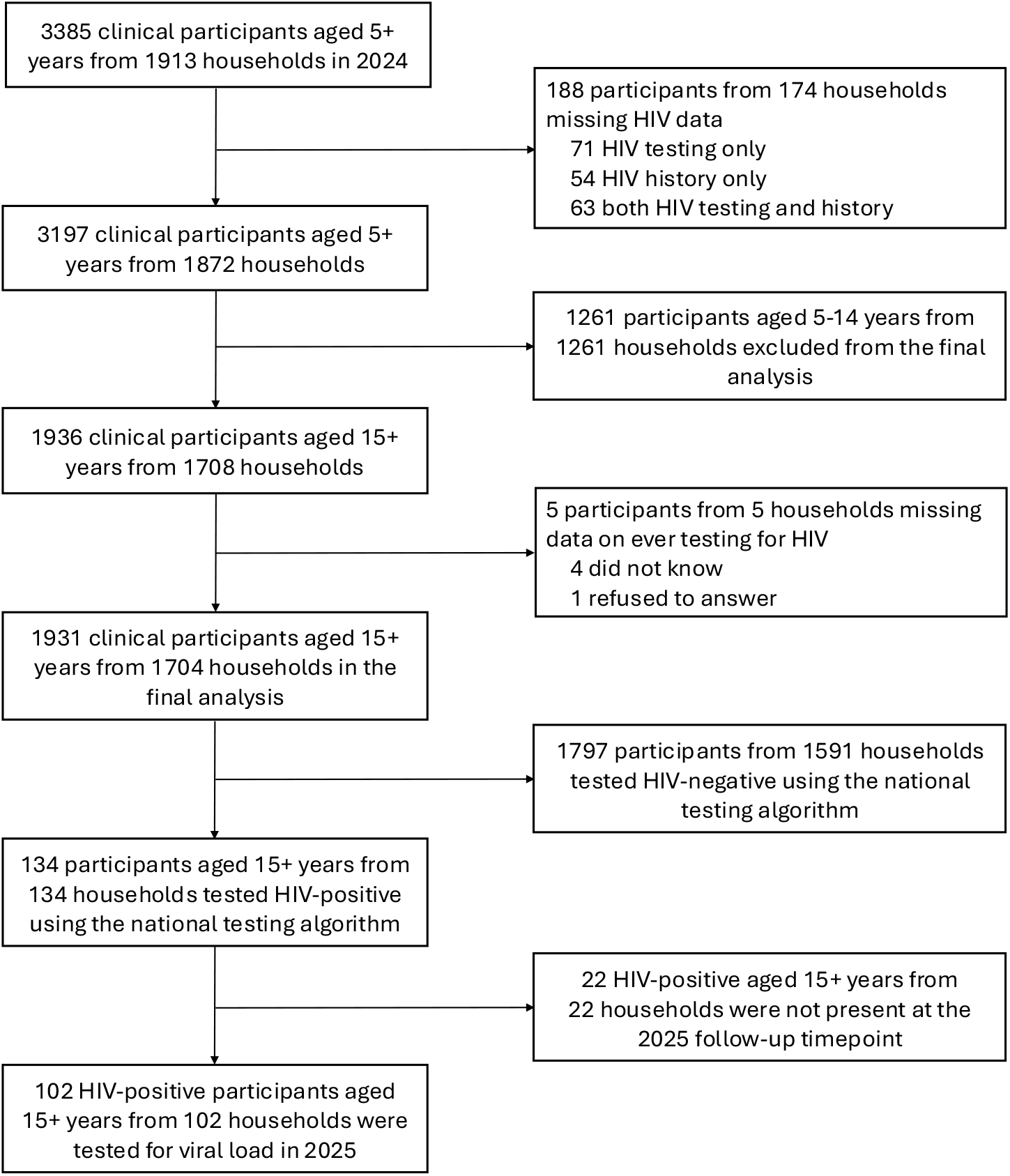
Participant flow diagram.

Samples for viral load quantification were collected for a subset of PLHIV who attended the follow-up event in January 2025 (124 out of 163 eligible by HIV positivity on Determine). Viral load was collected and analysed using dried blood spots (m2000sp) and viral suppression was defined as a binary indicator of a viral load result of <1000 copies/mL, in line with WHO criteria [5, 24]. Detailed information on HIV data collection is provided in the Supplementary Methods.

### Fisherfolk definitions

The household survey was conducted at recruitment, with participants enrolled between 2022 and 2024. It was administered by local surveyors fluent in Lusoga, Lugungu, Alur, and English. The survey collected demographic, socioeconomic, and health behavioural data, along with information on water-contact activities, household locations, and government health centre locations. Six definitions of fisherfolk were tested, three of which were individual-level and three were community-level. Two individual-level binary classifications were constructed based on participant reports of fishing activities (catching fish) and fishmongering activities (selling fish) among eleven water-contact activities, as detailed elsewhere [25]. We also examined a categorical occupation variable at the individual level, with categories including fisherman, fishmonger, subsistence farmer, and rice farmer, using none or other as the reference group. Community-level definitions were based on household distance to the nearest water site (continuous), the presence of a landing site (binary), and the presence of a beach in the village (binary).

This analysis also included individual- and household-level covariates covering sociodemographic, health behavioural, and spatial factors, as listed in the Supplementary Methods. Detailed definitions of the covariates are provided elsewhere [22].

### Statistical analysis

Data were collected through digital surveys created using Open Data Kit Collect (versions 2022.4.2, 2023.2.4, and 2024.3), with data entered using Android tablets (Android 9.0 Pie software). Analyses were performed using R v4.2.1 [26].

Geographic coordinates of reported HIV clinics were geocoded using the Google Maps Geocoding Application Programming Interface (API) and mapped to Uganda district shapefiles [27, 28]. Geodesic distances were calculated between households and HIV clinics [29].

Descriptive analyses of HIV testing and status trends were conducted by constructing kernel density plots over age, stratified by gender, for all participants. Each density curve represented a smoothed estimate of the underlying age distribution for the corresponding outcome, calculated using a Gaussian kernel with the default bandwidth (Silverman’s rule of thumb) [26]. Only participants aged 15 years or older (referred to as adults for reporting purposes, even though this age group includes older adolescents) were included in the further analyses in order to be comparable with UPHIA national statistics [30]. Since the age distribution of the SchistoTrack population sample is bimodal, study HIV prevalence estimates were directly age-standardised to the UPHIA 2020-2021 population age structure for comparison with national HIV prevalence estimates. For this, participants were grouped into 10-year age strata, with those aged 65 and above grouped together.

To better understand the social, behavioural, and spatial factors associated with HIV testing and HIV infection status, we constructed adjusted logistic regression models. The models were specified through backwards stepwise model selection using the lowest Bayesian information criterion (BIC) [26]. The candidate covariate list was guided by determinants found relevant in published literature and included the six definitions of fisherfolk and other covariates detailed above [8, 31, 32]. Age, gender and district were forced into the models. Additionally, fisherfolk definitions were assessed using unadjusted and minimally adjusted logistic regression models (controlling for age, gender and district) to identify the most relevant definition based on associations with ever testing, testing in the past 12 months, and HIV status. One participant was missing information on tribal membership and another on alcohol and smoking; these two participants were excluded during stepwise model selection but included in the final models, as those variables were not retained. 95% confidence intervals for odds ratios in BIC-based models were derived from standard errors clustered at the village level, to account for the sampling design of households nested within villages [33]. Floating absolute risks were also calculated for the district variable to assess the chosen reference category [34]. Intraclass correlation coefficients (ICCs) for empty multilevel models at the village level were calculated to assess variations within and between clusters for the study outcomes [35]. Variance inflation factors (VIFs) were computed to assess multicollinearity [36]. Although the main objective of this analysis was inference rather than prediction, we provided a measure of predictive performance by calculating the mean area under the curve (AUC) of the receiver operating characteristic (ROC) curve over stratified ten-fold cross-validation [37]. Sensitivity analyses were conducted by re-running the logistic regression models to include participants who did not know their HIV testing history (ever testing or testing in the past 12 months) as non-tested. Additionally, as fishing activities were predominantly reported by males, a descriptive subgroup analysis of the main outcomes was performed among male adults only.

### Patient and public involvement

HIV district focal people, community members, adult study participants, and the Ministry of Health leaders were involved in the study design, analysis, and interpretation of results.

## RESULTS

### HIV prevalence and testing

A total of 3395 participants aged 5 years and older from 1913 households were examined in 2024 (Figure 1). 188 participants did not have HIV testing or history data available, leaving 3197 clinical participants from 1873 households. 124 participants who tested positive for HIV by Determine were tested for viral load in 2025. Study participant characteristics and HIV prevalence by subgroups are summarised in Table 1 and Supplementary Tables S2-S5. HIV prevalence in the entire SchistoTrack cohort was 4.66% (149/3197), including 1.19% (15/1261) in children younger than 15 years. Proportion and kernel density plots of ever testing, testing recency, and HIV positivity, stratified by age and gender, are shown in Figure 2. Subsequent analyses focus on adults aged 15 years and older, comprising 1931 participants from 1704 households (Figure 1). HIV prevalence among this group was 6.94% (134/1931), with an age-standardised prevalence of 5.58%. 21.49% (415/1931) of participants reported fishing activities, among whom HIV prevalence was 6.02% (25/415) and age-standardised prevalence was 3.93%. By district, HIV prevalence was 5.84% (27/462) in Mayuge, 7.17% (60/837) in Pakwach, and 7.44% (47/632) in Buliisa (χ^2^ = 1.17, *p* = 0.56). HIV prevalence in females was 8.21% (97/1181) and 4.93% (37/750) in males (χ^2^ = 7.14, *p* = 0.01).

**Table 1.**
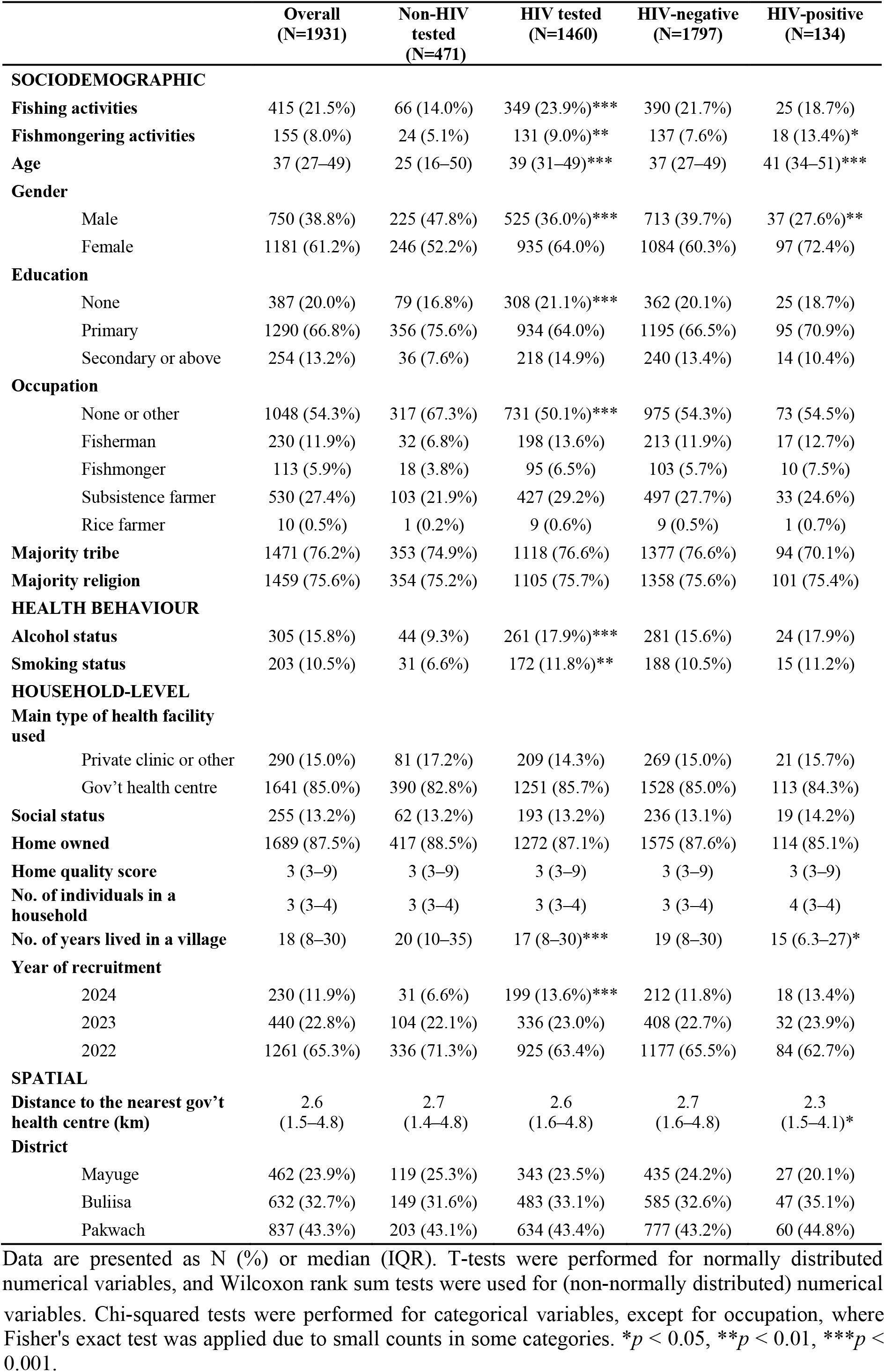
Adult participant characteristics (aged 15 and older), stratified by ever testing for HIV and HIV infection status.

**Figure 2.**
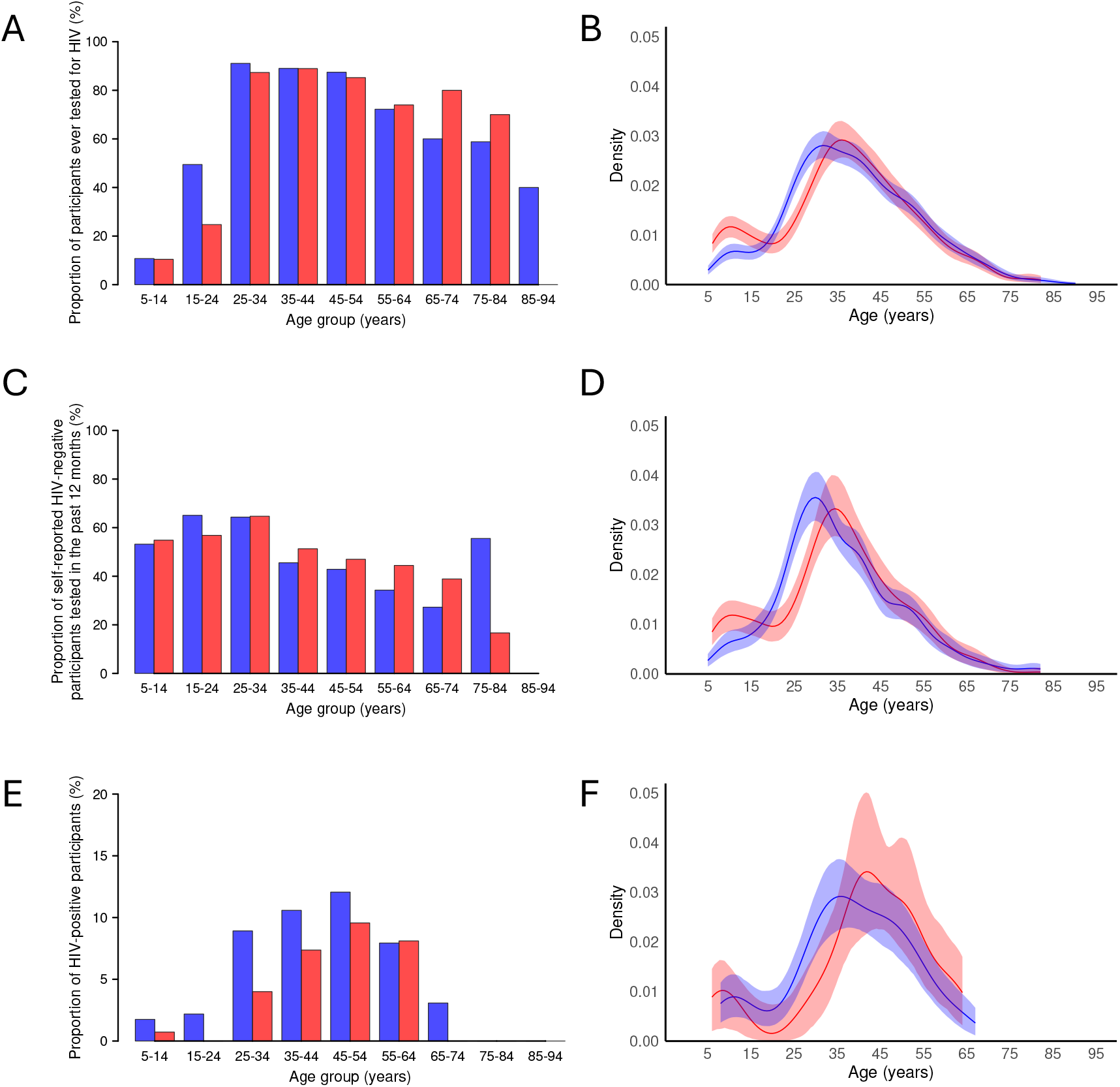
HIV testing coverage and prevalence by age and gender. (A) Proportion of participants ever tested for HIV (obs. = 3197). (B) Kernel density plot of ever testing. (C) Proportion of self-reported HIV-negative participants tested for HIV in the past 12 months (obs. = 1452). (D) Kernel density plot of testing in the past 12 months. (E) Proportion of HIV-positive participants (obs. = 3197). (F) Kernel density plot of HIV positivity. Males – red, females – blue.

75.61% (1460/1931) of participants reported previously undertaking an HIV test. There were no significant differences between districts for individuals ever tested for HIV (χ^2^ = 0.70, *p* = 0.70). 68.93% (1331/1931) of participants reported never receiving a positive HIV test result. Of those, 3.68% (49/1331) did not know when they last tested for HIV, with the majority (85.71%, 42/49) being from Pakwach. Approximately half of self-reported HIV-negative participants who recalled when they last tested for HIV had been tested in the past 12 months (51.87%, 665/1282). Similarly, about half of those reporting fishing activities had tested in the past 12 months (52.23%, 164/314), with no significant difference between the groups (χ^2^ = 0.01, *p* = 0.94). 65.44% (284/434) of participants in Buliisa were tested for HIV in the past 12 months, compared to only 49.03% (152/310) in Mayuge and 42.57% (229/538) in Pakwach (χ^2^ = 51.66, *p* < 0.001). 51.29% (418/815) of females and 52.89% (247/467) of males were tested in the past 12 months (χ^2^ = 0.24, *p* = 0.62). Detailed data on HIV testing recency stratified by fishing activities are provided in Figure S2.

22.39% (30/134) of PLHIV were newly diagnosed within the SchistoTrack study, of whom 16.67% (5/30) reported fishing activities. Only four newly diagnosed individuals reported never having tested for HIV prior to the study. Among status-aware PLHIV, 5.77% (6/104) had been first diagnosed within the previous 12 months (none of whom reported fishing activities), 47.12% (49/104) 1 to 9 years ago, and 47.12% (49/104) 10 or more years ago (Figure S2). Most status-aware PLHIV had been diagnosed at a health clinic or hospital (93.27%, 97/104); only 4.81% (5/104) had been diagnosed through outreach mobile testing, and only one person reported being diagnosed at an HIV-specific testing and counselling centre. More status-aware PLHIV from Mayuge (20%, 4/20) had been diagnosed through outreach mobile testing than from Buliisa (0%, 0/38) and Pakwach (2.17%, 1/46) (Fisher’s *p* = 0.005). Detailed information on testing locations stratified by fishing activities is shown in Figure S3.

### HIV care cascade

Among PLHIV reporting fishing activities, 80% (95% CI 64.32%-95.68%, 20/25) were aware of their HIV status, and 76% (95% CI 59.26%-92.74%, 19/25) were currently on ART. Self-reported viral suppression in this group was 100% (8/8) among PLHIV who had previously tested for viral load and knew the result. However, only 44% (11/25) of PLHIV reporting fishing activities recalled previously testing for viral load, and 32% (8/25) knew the result. Overall, 72% (95% CI 54.40%-89.60%, 18/25) of all PLHIV reporting fishing activities were linked to care, 80% (95% CI 64.32%-95.68%, 20/25) had initiated ART, and 76% (95% CI 59.26%-92.74%, 19/25) were retained in care (Figure S4). There were no significant differences in HIV care engagement between PLHIV reporting fishing activities and the general population or between districts (Figure S5). The results remained non-significant when the care cascade was re-examined using alternative fisherfolk definitions, including occupation (fishing or fishmongering) and fishmongering activities.

Overall, 88.06% (118/134) of PLHIV reported the HIV clinic where they accessed ART. Geographic coordinates were available for 86.96% (20/23) of the reported HIV clinics and 85.82% (115/134) of PLHIV. The median distance travelled to the reported HIV clinic was 6.73 kilometres (km) (IQR 2.64-11.61), compared to a median distance of 5.44 km (IQR: 2.28-7.58) to the nearest HIV clinic (Wilcoxon W = 561, *p* < 0.001). Participants from Pakwach travelled shorter distances to their reported HIV clinics (median 2.45 km, IQR 1.76-5.88) than those from Buliisa (6.95 km, IQR 5.37-13.46) and Mayuge (11.61 km, IQR 11.00-12.45) (Kruskal-Wallis χ^2^ = 44.86, *p* < 0.001). There were no significant differences by fishing activities (Kruskal-Wallis χ^2^ = 1.74, *p* = 0.19). 71.30% (82/115) of PLHIV accessed ART at a clinic other than their closest facility, with no significant differences by fishing activities (χ^2^ = 7.43 × 10^−31^, *p* = 1.00). However, there were district differences: 89.13% (41/46) of PLHIV from Pakwach used their nearest clinic, compared to 57.69% (15/26) from Mayuge and 60.47% (26/43) of PLHIV from Buliisa (χ^2^ = 11.97, *p* = 0.003). Those who did not access ART at their nearest clinic travelled a median distance of 10.59 km (IQR 4.07-34.99) farther than necessary (i.e., beyond the distance of the nearest clinic). Of those, PLHIV from Mayuge travelled a median additional distance of 1.18 km (IQR 0.79-10.33), compared to 34.26 km (IQR 6.86-39.90) in Buliisa and 21.03 km (IQR 12.58-34.94) in Pakwach (Kruskal-Wallis χ^2^ = 12.09, *p* = 0.002). Overall, 17.39% (20/115) of PLHIV received care outside their home district, with no significant differences by fishing activities or district (Fisher’s *p* = 0.24 and *p* = 0.06, respectively).

Among adult PLHIV aged 15 and older, 76.12% (102/134), including 68% (17/25) of PLHIV reporting fishing activities, had their viral load measured in 2025 (Figure 1). Among adult PLHIV with viral load, the median viral load was <839 copies/mL (limit of detection) (IQR <839-1124). 51.96% (53/102) of PLHIV had viral load below the limit of detection, while the viral load among those with detectable levels ranged from <839 to 203423 copies/mL. 70.59% (72/102) of PLHIV were virally suppressed (<1000 copies/mL). Among unsuppressed PLHIV, 30% (9/30) had a low-level unsuppressed viral load between 1000 and 2000 copies/mL. Viral suppression among PLHIV reporting fishing activities (70.59%, 12/17) was comparable to that of the general population (70.59%, 60/85), with no significant differences between districts: 81.82% (18/22) in Mayuge, 67.65% (23/34) in Buliisa, and 67.39% (31/46) in Pakwach. Among those newly diagnosed in 2024, the median viral load was <839 copies/mL (IQR <839-2980), with 61.90% (13/21) virally suppressed a year later. Of those unsuppressed, 25% (2/8) had a viral load between 1000 and 2000 copies/mL. Over half of PLHIV with measured viral load (64.71%, 66/102) reported previously testing for viral load and roughly half (48.04%, 49/102) knew the result. Agreement between self-reported viral load and measured viral suppression a year later was 79.59% (95% CI 65.66%-89.76%), with discordance driven by individuals reporting viral suppression when they were not actually suppressed (sensitivity 100%, 95% CI 90.51%-100%; specificity 16.67%, 95% CI 2.09%-48.41%).

### Associations between fishing activities and HIV status and testing

Of the six fisherfolk definitions tested, community-level definitions of a presence of a landing site in a village and household distance to the nearest water site were uninformative for the main outcomes of ever testing, testing in the past 12 months, and HIV status (Table 2). Presence of a beach in the village was positively associated with HIV status in unadjusted and minimally adjusted models but not selected in the BIC-based model. Fishmongering activities and occupation were crudely associated with ever testing, while only fishmongering activities were crudely associated with HIV status, but no fishmongering variables remained significant after adjustment for gender. Fishing occupation was associated with ever testing in unadjusted and minimally adjusted models but not selected in the BIC-based model. Only fishing activities remained robust for ever testing across unadjusted, minimally adjusted, and BIC-based models. No definitions of fisherfolk were associated with recent testing in unadjusted or minimally adjusted models.

**Table 2.** Association of fisherfolk definitions with HIV testing and status. Evaluation of six fisherfolk definitions using unadjusted and minimally adjusted (for age, gender and district) logistic regression models. The main outcomes were ever testing for HIV, testing in the past 12 months and HIV status. Analyses were conducted among all study participants (n = 1931) and among self-reported HIV-negative participants for the testing recency outcome (n = 1282). Only fisherman and fishmonger categories are shown for the occupation variable, which also included subsistence farmers and rice farmers.

| Outcome | Predictor | Unadjusted OR (95% CI) | Adjusted OR (95% CI) |
| --- | --- | --- | --- |
| Ever testing for HIV | Fishing activities | 1.93 (1.46–2.59)*** | 3.40 (2.45–4.77)*** |
|  | Fishmongering activities | 1.84 (1.19–2.94)** | 1.38 (0.88–2.25) |
|  | Occupation<br>(Ref. None or other) |  |  |
|  | Fisherman | 2.68 (1.83–4.05)*** | 3.53 (2.33–5.47)*** |
|  | Fishmonger | 2.29 (1.39–3.97)** | 1.65 (0.98–2.91) |
|  | Landing site | 0.89 (0.68–1.16) | 0.87 (0.66–1.14) |
|  | Beach | 1.14 (0.93–1.41) | 1.21 (0.92–1.59) |
|  | Household distance to water site (m) | 1.00 (1.00–1.00) | 1.00 (1.00–1.00) |
| Testing in the past 12 months | Fishing activities | 1.02 (0.79–1.32) | 0.86 (0.61–1.22) |
|  | Fishmongering activities | 1.10 (0.75–1.64) | 1.02 (0.67–1.58) |
|  | Occupation<br>(Ref. None or other) |  |  |
|  | Fisherman | 0.94 (0.67–1.32) | 0.86 (0.58–1.28) |
|  | Fishmonger | 1.27 (0.80–2.04) | 1.05 (0.64–1.73) |
|  | Landing site | 0.96 (0.73–1.27) | 0.88 (0.65–1.17) |
|  | Beach | 1.91 (1.53–2.39) | 1.44 (1.08–1.93) |
|  | Household distance to water site (m) | 1.00 (1.00–1.00) | 1.00 (1.00–1.00) |
| HIV infection status | Fishing activities | 0.83 (0.52–1.27) | 1.25 (0.70–2.22) |
|  | Fishmongering activities | 1.88 (1.08–3.11)* | 1.49 (0.83–2.55) |
|  | Occupation<br>(Ref. None or other) |  |  |
|  | Fisherman | 1.07 (0.60–1.80) | 1.75 (0.89–3.40) |
|  | Fishmonger | 1.30 (0.61–2.48) | 1.07 (0.50–2.09) |
|  | Landing site | 0.89 (0.59–1.38) | 0.87 (0.57–1.36) |
|  | Beach | 1.54 (1.07–2.23)* | 2.02 (1.28–3.23)** |
|  | Household distance to nearest water site (m) | 1.00 (1.00–1.00) | 1.00 (1.00–1.00) |

OR = odds ratio; CI = confidence interval. \* $p < 0.05$ , \*\* $p < 0.01$ , \*\*\* $p < 0.001$ .

The BIC-based model of ever testing for HIV included age, age^2^, gender, fishing activities, and district (Figure 3A). Older age was associated with higher odds of ever testing for HIV from 15 to 30 years of age (OR 1.32, 95% CI 1.25-1.40). Beyond this age range, the effect of age was not relevant. Including the quadratic term for age resulted in an attenuated odds ratio compared to the minimally adjusted model. Only 58.14% (357/614) of participants aged 15-30 had ever tested for HIV, compared to 83.75% (1103/1317) of participants aged over 30 years old. Female participants had 1.85 times higher odds of ever testing for HIV compared to male participants (CI 1.41-2.43). The proportion of ever tested participants was 79.17% (935/1181) in females and 70% (525/750) in males (χ^2^ = 20.42, *p* < 0.001). Participants reporting fishing activities had 1.76 times higher odds of ever testing compared to the general population (95% CI 1.22-2.54). 84.10% (349/415) of those reporting fishing activities and 73.28% (1111/1516) of the general population were ever tested for HIV (χ^2^ = 20.07, *p <* 0.001). Fishing activities were highly gendered: 91.33% (379/415) of participants reporting fishing activities were male. There were no consistent district effects across the models. The results remained robust after including participants who did not know their HIV testing history as non-tested and when analyses were restricted to male participants only (Figure S6 and Table S7).

**Figure 3.**
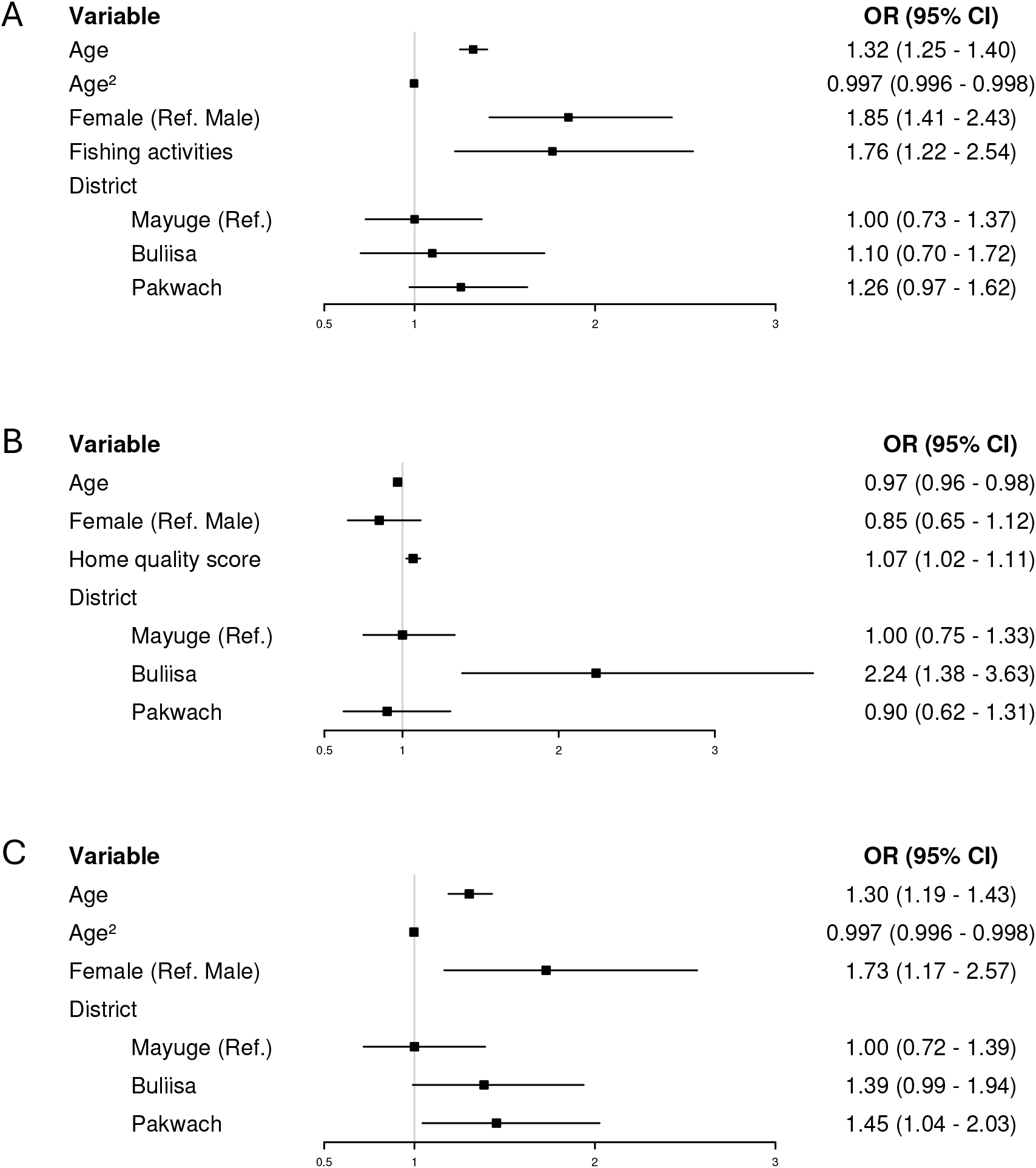
HIV testing and status models. (A) Model of ever testing for HIV (obs. = 1931). (B) Model of testing for HIV in the past 12 months (obs. = 1282). (C) Model of HIV status (obs. = 1931). Logistic regression models selected by backward stepwise selection based on the lowest Bayesian Information Criterion. 95% confidence intervals were calculated using village-level clustered standard errors (number of village clusters = 52). Floating absolute risks were calculated for the district variable. ICC = 0.041 (ever testing); 0.23 (testing recency); 0.024 (HIV infection). VIF range: 1.07-25.41 (ever testing); 1.03-1.43 (testing recency); 1.02-34.43 (HIV infection). AUC for stratified ten-fold cross-validation = 0.75 (ever testing); 0.66 (testing recency); 0.67 (HIV infection).

## DISCUSSION

Fishing communities in Uganda have long been recognised as a priority population for the HIV response, yet they remain underrepresented in national HIV surveillance data [9]. We screened 3197 individuals aged 5-92 years for HIV in the SchistoTrack cohort in rural Eastern and Western Uganda. We analysed HIV prevalence, testing coverage, and progress along the HIV care cascade in 1931 adults comprising the highest burden age group (aged 15 years and older). Our findings indicate that, in contrast to precedents, HIV prevalence in participants reporting fishing activities was not significantly higher than that of the general rural population. Participants who reported fishing activities reported higher lifetime HIV testing coverage and comparable care cascade engagement, although still fell short of current UNAIDS targets.

The age-standardised HIV prevalence in SchistoTrack (5.6%) closely mirrors the national prevalence reported by UPHIA 2020-2021 (5.8%) [30]. The moderately higher crude prevalence (6.8%) reflects the older age distribution of the SchistoTrack cohort compared to UPHIA. Over one-fifth (22.4%) of PLHIV in our cohort were newly diagnosed. Nearly three quarters (73.3%) of previously undiagnosed HIV cases were among women aged 25-52, which contrasts with broader trends across SSA, where women generally have better access to and uptake of HIV testing and care [38]. However, this may reflect the higher overall HIV burden among women or potential gender biases in self-reported data [1, 39].

In our study, participants reporting fishing activities did not have higher HIV prevalence than the general population. Contrary to earlier HIV prevalence estimates in Ugandan fishing communities between 15-40%, our study found a much lower age-standardised prevalence of 3.9% and crude prevalence of 6.0% in participants reporting fishing activities [7-9]. This may reflect our study locations being around the River Nile and Lake Albert, as opposed to Lake Victoria where most previous studies of Ugandan fishing communities have taken place and where HIV prevalence is high [8, 10, 11, 19, 20, 40]. Additionally, differences in sampling methodology (recruitment from landing sites), exclusion of older adults, and lack of age standardisation in other studies limit their generalisability and comparability [10-12, 19, 20]. Being fisherfolk was not relevant for identifying PLHIV at the individual level. Importantly, the lack of association between fishing activities and HIV status may also reflect the gendered nature of fishing: 91.3% of those reporting fishing activities were male, while the majority of PLHIV in our study were female. While the presence of a beach in the village was associated with increased odds of HIV in unadjusted and minimally adjusted models, this was not retained after variable selection, suggesting that community-level relevance of fisherfolk definitions needs additional exploration in studies with more diverse communities than studied here. The suggested importance of distinguishing between individual and community-level definitions of fisherfolk also warrants further investigation to accurately define priority groups and efficiently use limited ART resources. There were no significant differences in HIV prevalence between districts, and prevalences were comparable to regional UPHIA estimates.

78% of PLHIV were already aware of their status, of whom 97% reported being on ART, and among those who knew their viral load result, 93% reported viral suppression. This is similar to the 81-96-92 cascade reported by UPHIA, where status awareness represents a key challenge to achieving the UNAIDS 95-95-95 targets [30]. Among PLHIV reporting fishing activities, 80% were aware of their status, of whom 95% reported being on ART, and among those who knew their viral load result, all self-reported viral suppression. Despite this seemingly encouraging comparison, these results should be interpreted with caution, since many participants had either not tested for viral load due to being status-unaware or did not know their viral load result. The care cascade among PLHIV reporting fishing activities was close to that observed by Burgos-Soto et al. [12] in fishing communities around Lakes Edward and George (86-99-87). As in that study, care engagement among PLHIV reporting fishing activities was comparable to that of the general rural population, contributing to growing evidence that challenges previous expectations of poorer outcomes in this priority population. Yet these findings potentially indicate the success of targeted HIV care engagement interventions among fisherfolk in Uganda, who have historically been defined as a priority population [8, 9].

Measured viral suppression, available for a subset of participants who tested HIV-positive a year previously, was comparable between all adult PLHIV and those reporting fishing activities (70.6%), but was considerably lower than the national target of 85.7% among all PLHIV [17]. Low viral suppression (33.3%) among children, adolescents and young adults aligns with patterns commonly observed in SSA, as it is when new HIV diagnoses typically occur [1]. Only 61.9% of newly diagnosed adult PLHIV were virally suppressed one year after diagnosis, despite being linked to care and initiating ART. While 30% of those unsuppressed had relatively low viral load (1000-2000 copies/mL), suggesting possible measurement error, eight PLHIV had high viral loads (>10000 copies/mL) despite ART enrolment. Poor ART adherence (rather than treatment resistance) was the main challenge in this cohort, based on post-study follow-up of unsuppressed participants by the district HIV counsellors. The agreement between self-reported and measured viral load a year later was 79.6%, demonstrating that self-reported data can serve as a reliable proxy for viral suppression in the absence of laboratory results.

This agreement rose to 81.6% when the low but unsuppressed individuals were considered as suppressed. Notably, discordance was mainly driven by participants who self-reported viral suppression but were not suppressed in the following year. Future studies should examine whether this reflects misconceptions about viral suppression, actual treatment failure, or unreported adherence issues. Moreover, examining ART regimens through health records and assessing potential adherence challenges could help contextualise viral load findings. Many PLHIV (71.3%) in our study accessed ART at clinics farther from their nearest facility. Additional research is needed to understand whether individuals chose to travel longer distances due to concerns about stigma, heterogeneous healthcare infrastructure, or because they selected services closer to their workplace or fishing locations. If this behaviour reflects an active individual choice, it underscores the need for more accessible service delivery that accommodates personal preferences and needs.

Although most HIV outcomes analysed did not differ between those reporting fishing activities and the general population, these communities fell short of UNAIDS targets for care of PLHIV. Additionally, 24% of SchistoTrack participants had never tested for HIV and 48% had not tested in over a year, estimates comparable to UPHIA reports (22% and 57%, respectively) [30]. Individual-level definitions of fisherfolk were more relevant for ever testing than community-level definitions, further emphasising the need for more research into the importance of distinguishing between these levels. Fewer than 1% of previously tested individuals reported their last test or diagnosis was through HIV self-testing, revealing potential gaps in outreach efforts. Yet participants reporting fishing activities were more likely to have ever been tested for HIV, possibly reflecting prior targeted testing campaigns. Further work is needed to assess the apparently limited uptake of self-testing, given local knowledge (from district HIV counsellors) of past and current peer-based self-testing interventions in the study districts, and the literature indicating its suitability for reaching hard-to-reach, mobile fisherfolk [18, 41, 42]. The choice of testing modality may also affect different stages of the care cascade in distinct ways. For example, a comparison of home-based and outreach event-based HIV testing in Ugandan fishing communities around Lake Victoria found that home-based testing identified more undiagnosed cases but was linked to lower rates of linkage to care compared to event-based testing [43].

Our study has some limitations. Care cascade estimates were based on prevalent rather than follow-up data and relied solely on self-reports, potentially introducing memory error, recall and social desirability bias. The observed 100% ART uptake among those aware of their status may have been due to misreporting if participants were aware of their status primarily because of their ART use. Moreover, the twelve-month gap between self-reports and viral load measurement limits our ability to draw inference on the full care cascade, particularly if there is bias in who among PLHIV returned for follow-up. In addition, the relatively high limit of detection for DBS (839 copies/mL compared to typical thresholds of 50 or 200), lenient viral suppression threshold, and absence of cascade measure collection alongside the 2025 sample collection further limit interpretability. We also lacked detailed follow-up information on the exact ART regimen, individual adherence, and repeated viral load measures, which will be important in future studies to understand differences between viral load measurement error, treatment failure, and compliance issues. The relatively small number of PLHIV may have limited the statistical power to detect significant effects, potentially explaining the discrepancy between our findings and those of other studies reporting higher HIV prevalence (17.5%-41.3%) that have demonstrated significant differences [10-12, 20]. Importantly, this study should be replicated in other fishing community settings in Uganda and elsewhere in SSA to assess generalisability.

## CONCLUSION

Expanding community-based outreach, diversifying testing modalities, and addressing structural and mobility-related barriers remain critical to improving HIV status awareness and ensuring linkage to care. This study demonstrates the feasibility of leveraging established population-based studies, such as SchistoTrack, to generate representative estimates of HIV burden, prevention and care cascade engagement in under-served populations. We showed that individuals reporting fishing activities in rural Uganda have higher lifetime HIV testing rates and comparable HIV prevalence and care cascade engagement to the general population. These findings suggest that prior targeted outreach efforts may have contributed to improved testing uptake among fisherfolk. However, significant gaps persist in recent testing, status awareness, and viral suppression, underscoring the continued vulnerability of this priority population. To accelerate progress towards the UNAIDS 95-95-95 targets, it is crucial to continue to explore new strategies that promote regular HIV testing and sustained viral suppression.

## Data Availability

Participant data is not available due to the identifiable nature of the participant characteristics and ongoing nature of the cohort. Analysis code is provided as supplementary material.

## DECLARATIONS

## Acknowledgments

We thank the SchistoTrack Group for their valuable feedback. We also give most thanks to the field teams, including surveyors, technicians, nurses, and auxiliary workers. A special acknowledgment is to the study participants, village health team members, local government nurses and surveyors, clinical officers, and district leadership.

## Author contributions

GFC accepts full responsibility for the finished work and/or the conduct of the study, had access to the data, and controlled the decision to publish. Conceptualisation: GFC and ADS. Data curation: CP, BN, SWN, GWO, RM, JN, GFC. Formal analysis: HLB. Funding acquisition: GFC and ADS. Investigation: HLB, ADS, GFC. Methodology: LW, GFC. Project administration: NBK and GFC. Resources: NBK and GFC. Software: GFC. Supervision: GFC and ADS. Validation: HLB and GFC. Visualisation: HLB. Writing – original draft: HLB, GFC, and ADS. Writing – review & editing: HLB, LW, SWN, BN, RM, JN, GWO, CP, NBK, ADS, GFC.

## Ethics approvals

Data collection and use were reviewed and approved by Oxford Tropical Research Ethics Committee (OxTREC 509-21), the Vector Control Division Research Ethics Committee of the Uganda Ministry of Health (VCDREC146), and the Uganda National Council of Science and Technology (UNCST HS1664ES). Written informed consent was received from all adults, who also consented on behalf of those younger than 18 years. When possible, children also provided written informed consent. All children provided informed verbal assent.

