## Supplementary material for "HIV testing and prevalence in fishing communities in rural Uganda: a cross-sectional study of 3197 individuals within SchistoTrack"

### SUPPLEMENTARY METHODS

#### Adding HIV measures to SchistoTrack

##### I. Community engagement

In December 2023, study teams alongside neglected tropical disease (NTD) focal people (key public health personnel responsible for coordinating and overseeing the implementation of NTD-related activities at the district and sub-district levels) and HIV counsellors (local to the study districts) visited study villages and all adult participants in SchistoTrack were invited to attend community meetings. HIV counsellors led a session on HIV education and sensitisation to testing, addressing topics of stigma, viral suppression, the importance of testing, and the role of ART in living a long and healthy life. Community members were then asked to share their feelings about HIV testing within SchistoTrack and their view on how best to include testing in a way that suits individual households, community support mechanisms, access to ART, and, most importantly, maintenance of participant confidentiality. Participants shared that they would like the option of home self-test kits to take home to partners and for adults to be given their results without their children present, while adults were to be present when children were given results. It also was decided that ART initiation and coordination with care facilities will be done at the point-of-care for newly diagnosed participants (on the day of diagnosis) by the HIV counsellors, to avoid losing individuals to care and promptly start them on treatment. The counsellor also highlighted that HIV testing was optional and did not impact study inclusion; individuals were free to refuse testing. At the 2024 cohort timepoint all participants were reconsented to include HIV testing; written consent was obtained from all participants where adult participants consented on behalf of children, and children provided verbal assent after the adults.

##### II. Development of HIV history survey

An HIV diagnosis, treatment and care survey was adapted from established sources, including the WHO and UPHIA HIV surveys, as well as Uganda Ministry of Health HIV reports. It was also reviewed by officials from the Uganda Ministry of Health involved in national HIV programmes. SchistoTrack teams and HIV counsellors validated the HIV history survey during community engagement activities and training sessions, ensuring alignment with national guidelines and broader clinical consensus. The final survey comprised 26 questions covering prior HIV testing, diagnosis, treatment, access to care, and viral load testing (Table S1). No questions were asked about sexual partnerships, consistent with the wishes of study communities.

#### HIV survey and testing procedures

Participants visited an HIV counsellor who provided pre-test counselling and obtained written consent for new HIV study procedures. Consented participants completed an HIV history survey, which was conducted in private for adults while children were accompanied by an adult. HIV status was determined according to the Ministry of Health in Uganda serial testing algorithm based on three tests: Determine HIV-1/2 (Abbott) as a first-pass screening test, HIV 1/2 STAT-PAK Assay (Chembio Diagnostics) as a confirmatory test, and Bioline HIV 1/2 3.0 (Abbott) as a tie-breaker where the first two tests disagreed (Figure S1) [9]. Following the national algorithm, we considered the HIV test as positive if Determine and STAT-PAK were both positive, or if Determine was positive, STAT-PAK was negative, and Bioline was positive. The HIV counsellor-provided post-test counselling and HIV test results. The HIV clinical station in SchistoTrack was set up in a private room away from other clinical stations and ongoing cohort activities. To incentivise individuals to return to the HIV counsellor, participant compensation (a household-sized bar of soap) was collected from that station. Newly diagnosed HIV-positive participants were immediately linked to care where they received ART provision in line with national guidelines. Those requiring urgent medical attention were transported to local healthcare facilities.

##### Viral load quantification

Viral load was quantified using the Abbott RealTime HIV-1 assay in whole blood spotted on cards as dried blood spots (DBS), which had a detectable limit of 839 copies/mL (one-spot, 70 µL). One DBS card was assigned to each participant, and five blood spots per participant were taken. DBS samples were stored in -20°C for three months before processing. Samples were analysed across six runs in batches of twenty-one. All controls (negative, low HIV-positive and high HIV-positive) performed consistently across runs. The viral load range was 14995531-19025878 copies/mL for high HIV-positive controls and 79105-98836 for low HIV-positive controls. All negative controls had an undetected viral load. An internal control (HIV-unrelated sequence) was introduced into each sample to ensure correct sample processing. One participant had an invalid DBS result on the first run and required a second DBS sample to be processed.

##### Covariates used in the analysis

Individual-level covariates used in this analysis were age, gender, education, tribal membership, religion, current alcohol consumption, and current smoking. Household-level covariates were household size, home quality score, years of residence in the village, home ownership status, social status, year of recruitment, and the primary type of healthcare facility used. Spatial factors included the household distance from the nearest government health facility and district, which was categorised with Mayuge (East) as the reference level for comparison with Buliisa and Pakwach (West). The primary type of healthcare facility used was binary (government health facility with private or other facility serving as a reference category).

**Table S1.** SchistoTrack HIV survey questions used in the analysis.

| Variable | Question | Answer options |
|---|---|---|
| Ever testing for HIV | Have you ever tested for HIV? | 1. No<br>2. Yes<br>3. Do not know<br>4. Refused to answer |
| Self-reported HIV status (HIV status awareness) | Have you ever tested positive for HIV? | 1. No<br>2. Yes<br>3. Do not know<br>4. Refused to answer |
| When last negative HIV<br><br>(test HIV testing in the past 12 months among those who self-reported HIV-negative) | When did you last test negative for HIV? | 1. Within the last 3 months<br>2. Between 3 to 6 months ago<br>3. Between more than 6 to 12 months ago<br>4. More than 12 months ago<br>5. Do not know<br>5. Refused to answer |
| Linkage to care (within 30 days of diagnosis) | From the date that you learned you had HIV, how soon did you first see a healthcare provider for HIV care? | 1. Immediately, within a day<br>2. Between 1 to 7 days<br>3. Between 8 to 30 days<br>4. Between 31 days to 12 months<br>5. More than a year |
| Retention in care (access to care within the past 12 months) | When was the last time you received HIV care from a healthcare provider? | 1. Within the last 6 months<br>2. 6 to 12 months ago<br>3. More than 1 year ago<br>4. Do not know<br>5. Refused to answer |
| Initiation of ART | Have you ever taken ARVs, that is, antiretroviral medication to treat your HIV? | 1. No<br>2. Yes<br>3. Do not know<br>4. Refused to answer |
| Current use of ART | Are you currently taking ARVs to treat your HIV? | 1. No<br>2. Yes<br>3. Do not know<br>4. Refused to answer |
| Ever viral load | Some people with HIV get their viral load measured. This is a test that measures how much HIV is in your blood. Did you ever have a viral load test? | 1. No<br>2. Yes<br>3. Do not know<br>4. Refused to answer |
| Self-reported viral load result | What was the result of the last viral load test result? | 1. No virus detected<br>2. <200 copies/ml<br>3. 200-999 copies/ml<br>4. >=1000 copies/ml<br>5. Do not know<br>6. Refused to answer |
| When first positive HIV test | When did you first test positive for HIV? | 1. Within the last year<br>2. 1 to 4 years ago<br>3. 5 to 9 years ago<br>4. 10 to 14 years ago<br>5. >=15 years ago<br>6. Refused to answer |
| Where first positive test | Where did you have your first positive HIV test? | 1. Testing and counselling centre<br>2. Health clinic, hospital, or similar<br>3. Outreach mobile testing<br>4. Home self-test<br>5. Other, specify<br>6. Do not know<br>7. Refused to answer |
| Where last negative test | Where did you have your last negative HIV test? | 1. Testing and counselling centre<br>2. Health clinic, hospital, or similar<br>3. Outreach mobile testing<br>4. Home self-test<br>5. Other, specify<br>6. Do not know<br>7. Refused to answer |

**Table S2.** HIV prevalence by participant characteristics (adults 15 years and older).

| | Overall (N=1931) | HIV-positive (N=134) |
|---|---|---|
| **SOCIODEMOGRAPHIC FACTORS** | | |
| **Fishing activities** | | |
| Yes | 415 | 25 (6.0%) |
| No | 1516 | 109 (7.2%) |
| **Fishmongering activities** | | |
| Yes | 155 | 18 (11.6%) |
| No | 1776 | 116 (6.5%) |
| **Age group** | | |
| 15-24 | 368 | 4 (1.1%) |
| 25-49 | 1088 | 91 (8.4%) |
| 50+ | 475 | 39 (8.2%) |
| **Gender** | | |
| Male | 750 | 37 (4.9%) |
| Female | 1181 | 97 (8.2%) |
| **Education** | | |
| None | 387 | 25 (6.5%) |
| Primary | 1290 | 95 (7.4%) |
| Secondary or above | 254 | 14 (5.5%) |
| **Occupation** | | |
| None or other | 1048 | 73 (7.0%) |
| Fisherman | 230 | 17 (7.4%) |
| Fishmonger | 113 | 10 (8.8%) |
| Subsistence farmer | 530 | 33 (6.2%) |
| Rice farmer | 10 | 1 (10.0%) |
| **Majority tribe** | | |
| Yes | 1471 | 94 (6.4%) |
| No | 459 | 39 (8.5%) |
| **Majority religion** | | |
| Yes | 1459 | 101 (6.9%) |
| No | 472 | 33 (7.0%) |
| **HEALTH BEHAVIOURAL FACTORS** | | |
| **Alcohol status** | | |
| Yes | 305 | 24 (7.9%) |
| No | 1625 | 109 (6.7%) |
| **Smoking status** | | |
| Yes | 203 | 15 (7.4%) |
| No | 1727 | 118 (6.8%) |
| **HOUSEHOLD-LEVEL FACTORS** | | |
| **Main type of health facility used** | | |
| Private clinic or other | 290 | 21 (7.2%) |
| Gov't health centre | 1641 | 113 (6.9%) |
| **Social status** | | |
| Yes | 255 | 19 (7.5%) |
| No | 1676 | 115 (6.9%) |
| **Home owned** | | |
| Yes | 1689 | 114 (6.7%) |
| No | 242 | 20 (8.3%) |

**Table S2.** Continued.

| | Overall (N=1931) | HIV-positive (N=134) |
|---|---|---|
| **Home quality score** | | |
| 3 | 1016 | 70 (6.9%) |
| 4-8 | 367 | 25 (6.8%) |
| 9-12 | 548 | 39 (7.1%) |
| **No. of individuals in a household** | | |
| 2 | 350 | 30 (8.6%) |
| 3 | 693 | 35 (5.1%) |
| 4+ | 888 | 69 (7.8%) |
| **No. of years lived in a village** | | |
| 1-7 | 423 | 38 (9.0%) |
| 8-17 | 510 | 41 (8.0%) |
| 18-29 | 444 | 22 (5.0%) |
| 30+ | 554 | 33 (6.0%) |
| **Year of recruitment** | | |
| 2024 | 230 | 18 (7.8%) |
| 2023 | 440 | 32 (7.3%) |
| 2022 | 1261 | 84 (6.7%) |
| **SPATIAL FACTORS** | | |
| **Distance to the nearest gov't health centre (km)** | | |
| <=1.5 | 468 | 33 (7.1%) |
| ]1.5-2.5] | 431 | 42 (9.7%) |
| ]2.5-5.0] | 579 | 38 (6.6%) |
| >5.0 | 453 | 21 (4.6%) |
| **District** | | |
| Mayuge | 462 | 27 (5.8%) |
| Buliisa | 632 | 47 (7.4%) |
| Pakwach | 837 | 60 (7.2%) |

Data are presented as N (%).

**Table S3.** Participant characteristics stratified by HIV infection status (5 years and older).

| | **Overall (N=3197)** | **HIV-negative (N=3048)** | **HIV-positive (N=149)** | ***p*-value** |
|---|---|---|---|---|
| **SOCIODEMOGRAPHIC FACTORS** | | | | |
| **Fishing activities** | 438 (13.7%) | 413 (13.5%) | 25 (16.8%) | 0.001 |
| **Fishmongering activities** | 158 (4.9%) | 140 (4.6%) | 18 (12.1%) | |
| **Age** | 24 (11–41) | 20 (10–40) | 40 (32–50) | <0.001 |
| **Gender** | | | | <0.001 |
| Male | 1442 (45.1%) | 1400 (45.9%) | 42 (28.2%) | |
| Female | 1755 (54.9%) | 1648 (54.1%) | 107 (71.8%) | |
| **Education** | | | | 0.764 |
| None | 595 (18.6%) | 569 (18.7%) | 26 (17.4%) | |
| Primary | 2347 (73.4%) | 2238 (73.4%) | 109 (73.2%) | |
| Secondary or above | 255 (8.0%) | 241 (7.9%) | 14 (9.4%) | |
| **Occupation** | | | | 0.009 |
| None or other | 2300 (71.9%) | 2212 (72.6%) | 88 (59.1%) | |
| Fisherman | 231 (7.2%) | 214 (7.0%) | 17 (11.4%) | |
| Fishmonger | 113 (3.5%) | 103 (3.4%) | 10 (6.7%) | |
| Subsistence farmer | 542 (17.0%) | 509 (16.7%) | 33 (22.1%) | |
| Rice farmer | 11 (0.3%) | 10 (0.3%) | 1 (0.7%) | |
| **Majority tribe** | 2431 (76.0%) | 2325 (76.3%) | 106 (71.1%) | 0.213 |
| **Majority religion** | 2416 (75.6%) | 2301 (75.5%) | 115 (77.2%) | 0.777 |
| **HEALTH BEHAVIOURAL FACTORS** | | | | |
| **Alcohol status** | 307 (9.6%) | 283 (9.3%) | 24 (16.1%) | 0.007 |
| **Smoking status** | 206 (6.4%) | 191 (6.3%) | 15 (10.1%) | 0.084 |
| **Main type of health facility used** | | | | 1.000 |
| Private clinic or other | | | | |
| Government health centre | 473 (14.8%) | 451 (14.8%) | 22 (14.8%) | |
| **HOUSEHOLD-LEVEL FACTORS** | 2724 (85.2%) | 2597 (85.2%) | 127 (85.2%) | |
| **Social status** | 410 (12.8%) | 391 (12.8%) | 19 (12.8%) | 1.000 |
| **Home owned** | 2800 (87.6%) | 2673 (87.7%) | 127 (85.2%) | 0.560 |
| **Home quality score** | 3 (3–9) | 3 (3–9) | 3 (3–9) | 0.770 |
| **No. of individuals in a household** | 3 (3–4) | 3 (3–4) | 4 (3–4) | 0.797 |
| **No. of years lived in a village** | 18 (8–30) | 18 (8–30) | 14 (6–27) | 0.013 |
| **Year of recruitment** | | | | 0.996 |
| 2024 | 441 (13.8%) | 421 (13.8%) | 20 (13.4%) | |
| 2023 | 733 (22.9%) | 697 (22.9%) | 36 (24.2%) | |
| 2022 | 2023 (63.3%) | 1930 (63.3%) | 93 (62.4%) | |
| **SPATIAL FACTORS** | | | | |
| **Distance to the nearest gov't health centre (km)** | 2.6 (1.5–4.8) | 2.6 (1.5–4.8) | 2.4 (1.53–4.2) | 0.099 |
| **District** | | | | 0.572 |
| Mayuge | 753 (23.6%) | 723 (23.7%) | 30 (20.1%) | |
| Buliisa | 1073 (33.6%) | 1018 (33.4%) | 55 (36.9%) | |
| Pakwach | 1371 (42.9%) | 1307 (42.9%) | 64 (43.0%) | |

Data are presented as N (%) or median (IQR). T-tests were performed for normally distributed numerical variables, and Wilcoxon rank sum tests were used for (non-normally distributed) numerical variables. Chi-square tests were performed for categorical variables, except for occupation, where Fisher's exact test was applied due to small counts in some categories.

**Table S4.** Participant characteristics stratified by testing in the past 12 months (self-reported HIV-negative adults 15 years and older).

| | Overall (N=1282) | Non-HIV tested (N=617) | HIV tested (N=665) | *p*-value |
|---|---|---|---|---|
| **SOCIODEMOGRAPHIC FACTORS** | | | | |
| **Fishing activities** | 314 (24.5%) | 150 (24.3%) | 164 (24.7%) | 0.512 |
| **Fishmongering activities** | 109 (8.5%) | 50 (8.1%) | 59 (8.9%) | |
| **Age** | 38 (30–48) | 41 (32–51) | 35 (29–45) | <0.001 |
| **Gender** | | | | 0.836 |
| Male | 467 (36.4%) | 220 (35.7%) | 247 (37.1%) | |
| Female | 815 (63.6%) | 397 (64.3%) | 418 (62.9%) | |
| **Education** | | | | <0.001 |
| None | 270 (21.1%) | 146 (23.7%) | 124 (18.6%) | |
| Primary | 813 (63.4%) | 402 (65.2%) | 411 (61.8%) | |
| Secondary or above | 199 (15.5%) | 69 (11.2%) | 130 (19.5%) | |
| **Occupation** | | | | 0.001 |
| None or other | 643 (50.2%) | 290 (47.0%) | 353 (53.1%) | |
| Fisherman | 176 (13.7%) | 82 (13.3%) | 94 (14.1%) | |
| Fishmonger | 84 (6.6%) | 33 (5.3%) | 51 (7.7%) | |
| Subsistence farmer | 371 (28.9%) | 206 (33.4%) | 165 (24.8%) | |
| Rice farmer | 8 (0.6%) | 6 (1.0%) | 2 (0.3%) | |
| **Majority tribe** | 986 (76.9%) | 497 (80.6%) | 489 (73.5%) | 0.001 |
| **Majority religion** | 968 (75.5%) | 480 (77.8%) | 488 (73.4%) | 0.039 |
| **HEALTH BEHAVIOURAL FACTORS** | | | | |
| **Alcohol status** | 230 (17.9%) | 118 (19.1%) | 112 (16.8%) | 0.347 |
| **Smoking status** | 149 (11.6%) | 84 (13.6%) | 65 (9.8%) | 0.042 |
| **HOUSEHOLD-LEVEL FACTORS** | | | | |
| **Main type of health facility used** | | | | 0.367 |
| Private clinic or other | 178 (13.9%) | 90 (14.6%) | 88 (13.2%) | |
| Government health centre | 1104 (86.1%) | 527 (85.4%) | 577 (86.8%) | |
| **Social status** | 169 (13.2%) | 81 (13.1%) | 88 (13.2%) | 0.916 |
| **Home owned** | 1115 (87.0%) | 558 (90.4%) | 557 (83.8%) | <0.001 |
| **Home quality score** | 3 (3–9) | 3 (3–9) | 6 (3–9) | <0.001 |
| **No. of individuals in a household** | 3 (3–4) | 3 (3–4) | 3 (3–4) | 0.731 |
| **No. of years lived in a village** | 18 (8–30) | 20 (9–30) | 15 (8–26) | <0.001 |
| **Year of recruitment** | | | | 0.516 |
| 2024 | 176 (13.7%) | 78 (12.6%) | 98 (14.7%) | |
| 2023 | 300 (23.4%) | 147 (23.8%) | 153 (23.0%) | |
| 2022 | 806 (62.9%) | 392 (63.5%) | 414 (62.3%) | |
| **SPATIAL FACTORS** | | | | |
| **Distance to the nearest gov't health centre (km)** | 2.7 (1.6–4.8) | 2.7 (1.8–5.3) | 2.6 (1.2–4.7) | <0.001 |
| **District** | | | | <0.001 |
| Mayuge | 310 (24.2%) | 158 (25.6%) | 152 (22.9%) | |
| Buliisa | 434 (33.9%) | 150 (24.3%) | 284 (42.7%) | |
| Pakwach | 538 (42.0%) | 309 (50.1%) | 229 (34.4%) | |

Data are presented as N (%) or median (IQR). T-tests were performed for normally distributed numerical variables, and Wilcoxon rank sum tests were used for (non-normally distributed) numerical variables. Chi-square tests were performed for categorical variables, except for occupation, where Fisher's exact test was applied due to small counts in some categories.

**Table S5.** Participant characteristics stratified by fishing activities (adults 15 years and older).

| | Overall (N=1931) | Non-fishers (N=1516) | Fishers (N=415) | *p*-value |
|---|---|---|---|---|
| **SOCIODEMOGRAPHIC FACTORS** | | | | |
| **Age** | 37 (27–49) | 37 (26–50) | 38 (30.5–46) | 0.015 |
| **Gender** | | | | <0.001 |
| Male | 750 (38.8%) | 371 (24.5%) | 379 (91.3%) | |
| Female | 1181 (61.2%) | 1145 (75.5%) | 36 (8.7%) | |
| **Education** | | | | <0.001 |
| None | 387 (20.0%) | 371 (24.5%) | 16 (3.9%) | |
| Primary | 1290 (66.8%) | 973 (64.2%) | 317 (76.4%) | |
| Secondary or above | 254 (13.2%) | 172 (11.3%) | 82 (19.8%) | |
| **Occupation** | | | | <0.001 |
| None or other | 1048 (54.3%) | 917 (60.5%) | 131 (31.6%) | |
| Fisherman | 230 (11.9%) | 23 (1.5%) | 207 (49.9%) | |
| Fishmonger | 113 (5.9%) | 96 (6.3%) | 17 (4.1%) | |
| Subsistence farmer | 530 (27.4%) | 472 (31.1%) | 58 (14.0%) | |
| Rice farmer | 10 (0.5%) | 8 (0.5%) | 2 (0.5%) | |
| **Majority tribe** | 1471 (76.2%) | 1111 (73.3%) | 360 (86.7%) | <0.001 |
| **Majority religion** | 1459 (75.6%) | 1130 (74.5%) | 329 (79.3%) | 0.080 |
| **HEALTH BEHAVIOURAL FACTORS** | | | | |
| **Alcohol status** | 305 (15.8%) | 135 (8.9%) | 170 (41.0%) | <0.001 |
| **Smoking status** | 203 (10.5%) | 79 (5.2%) | 124 (29.9%) | <0.001 |
| **HOUSEHOLD-LEVEL FACTORS** | | | | |
| **Main type of health facility used** | | | | 0.015 |
| Private clinic or other | 290 (15.0%) | 231 (15.2%) | 59 (14.2%) | |
| Government health centre | 1641 (85.0%) | 1285 (84.8%) | 356 (85.8%) | |
| **Social status** | 255 (13.2%) | 183 (12.1%) | 72 (17.3%) | 0.029 |
| **Home owned** | 1689 (87.5%) | 1322 (87.2%) | 367 (88.4%) | 0.597 |
| **Home quality score** | 3 (3–9) | 3 (3–9) | 3 (3–6) | <0.001 |
| **No. of individuals in a household** | 3 (3–4) | 3 (3–4) | 3 (3–4) | 0.172 |
| **No. of years lived in a village** | 18 (8–30) | 18 (8–30) | 20 (9.5–30) | 0.811 |
| **Year of recruitment** | | | | <0.001 |
| 2024 | 230 (11.9%) | 175 (11.5%) | 55 (13.3%) | |
| 2023 | 440 (22.8%) | 337 (22.2%) | 103 (24.8%) | |
| 2022 | 1261 (65.3%) | 1004 (66.2%) | 257 (61.9%) | |
| **SPATIAL FACTORS** | | | | |
| **Distance to the nearest gov't health centre (km)** | 2.6 (1.5–4.8) | 2.6 (1.5–4.8) | 2.8 (1.6–4.8) | 0.477 |
| **District** | | | | <0.001 |
| Mayuge | 462 (23.9%) | 420 (27.7%) | 42 (10.1%) | |
| Buliisa | 632 (32.7%) | 490 (32.3%) | 142 (34.2%) | |
| Pakwach | 837 (43.3%) | 606 (40.0%) | 231 (55.7%) | |

Data are presented as N (%) or median (IQR). T-tests were performed for normally distributed numerical variables, and Wilcoxon rank sum tests were used for (non-normally distributed) numerical variables. Chi-square tests were performed for categorical variables, except for occupation, where Fisher's exact test was applied due to small counts in some categories.

**Table S6. Unadjusted associations of main outcomes.** Univariate logistic regression models of covariates selected by backward selection using the lowest BIC, assessing associations with ever testing for HIV and HIV status among all study participants (n = 1931), and testing in the past 12 months among self-reported HIV-negative participants (n = 1282).

| Outcome | Predictor | Reference level | Comparison level | OR (95% CI) | *p*-value |
|---|---|---|---|---|---|
| Ever testing for HIV | Age | | | 1.03 (1.02–1.04) | <0.001 |
| | $Age^2$ | | | 1.00 (1.00-1.00) | <0.001 |
| | Gender | Male | Female | 1.63 (1.32–2.01) | <0.001 |
| | Fishing activities | | | 1.93 (1.46–2.59) | <0.001 |
| | District | Mayuge | Buliisa | 1.12 (0.85–1.48) | 0.407 |
| | | | Pakwach | 1.08 (0.83–1.41) | 0.548 |
| Testing in the past 12 months | Age | | | 0.97 (0.96–0.98) | <0.001 |
| | Gender | Male | Female | 0.94 (0.75–1.18) | 0.581 |
| | Home quality | | | 1.05 (1.02–1.08) | <0.001 |
| | District | Mayuge | Buliisa | 1.97 (1.46–2.65) | <0.001 |
| | | | Pakwach | 0.77 (0.58–1.02) | 0.068 |
| HIV infection status | Age | | | 1.01 (1.00–1.03) | 0.008 |
| | $Age^2$ | | | 1.00 (1.00-1.00) | 0.169 |
| | Gender | Male | Female | 1.72 (1.18–2.58) | 0.006 |
| | District | Mayuge | Buliisa | 1.29 (0.80–2.14) | 0.301 |
| | | | Pakwach | 1.24 (0.79–2.02) | 0.361 |

OR = odds ratio; CI = confidence interval.

**Table S7. Summary of main outcomes among male adults (sensitivity analysis).** Adults were defined as participants aged 15 years and older. Obs. = 750 for HIV status and history of HIV testing; 467 for HIV testing in the past 12 months (among self-reported HIV-negative individuals); 37 PLHIV for the self-reported HIV care cascade, of whom 16 self-reported viral load results; 24 PLHIV had viral load measured. Chi-square tests were used for categorical variables, except for status awareness, current ART use, and measured viral suppression, where Fisher's exact test was applied due to small counts in some categories. Data are based on self-reports from the 2024 HIV survey and viral load measurements collected in 2025.

| | **Overall** | **Fishing activities** | **Other or no water activities** | ***p*-value** |
|---|---|---|---|---|
| HIV-positive | 4.9% (37/750) | 6.3% (24/379) | 3.5% (13/371) | 0.105 |
| Ever tested | 70.0% (525/750) | 84.2% (319/379) | 55.5% (206/371) | <0.001 |
| Tested in the past 12 months | 52.9% (247/467) | 52.8% (150/284) | 53.0% (97/183) | 1.000 |
| HIV care cascade | | | | |
| Status awareness | 78.4% (29/37) | 83.3% (20/24) | 69.2% (9/13) | 0.594 |
| Current ART | 93.1% (27/29) | 79.2% (19/20) | 61.5% (8/13) | 0.532 |
| Viral suppression (self-reported) | 100.0% (16/16) | 100.0% (8/8) | 100.0% (8/8) | 1.000 |
| Viral suppression (measured) | 70.8% (17/24) | 68.8% (11/16) | 75.0% (2/8) | 1.000 |

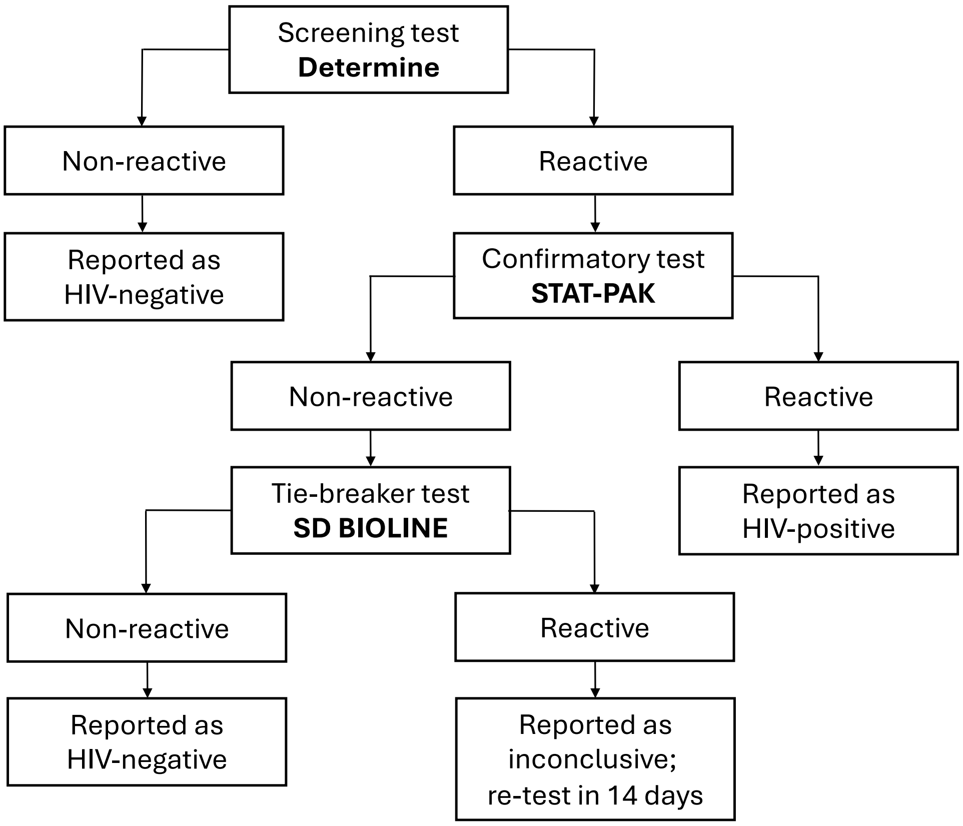

**Figure S1.** National HIV testing algorithm in Uganda.

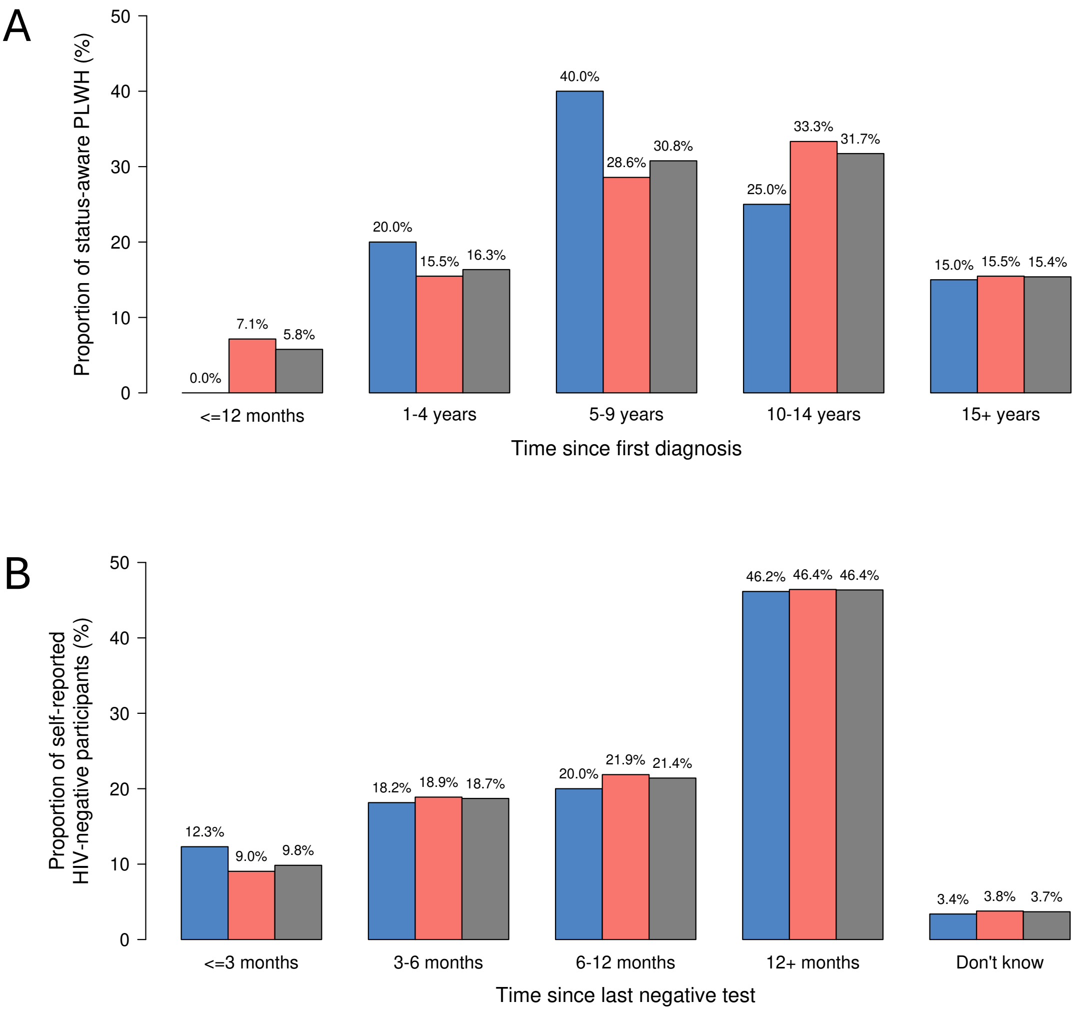

**Figure S2. Testing recency.** Time since first HIV diagnosis (status-aware participants) and time since last negative HIV test (self-reported HIV-negative participants), stratified by fishing activities (blue – fishing activities; red – no fishing activities; grey – overall). Data are based on participant self-reports from the HIV survey.

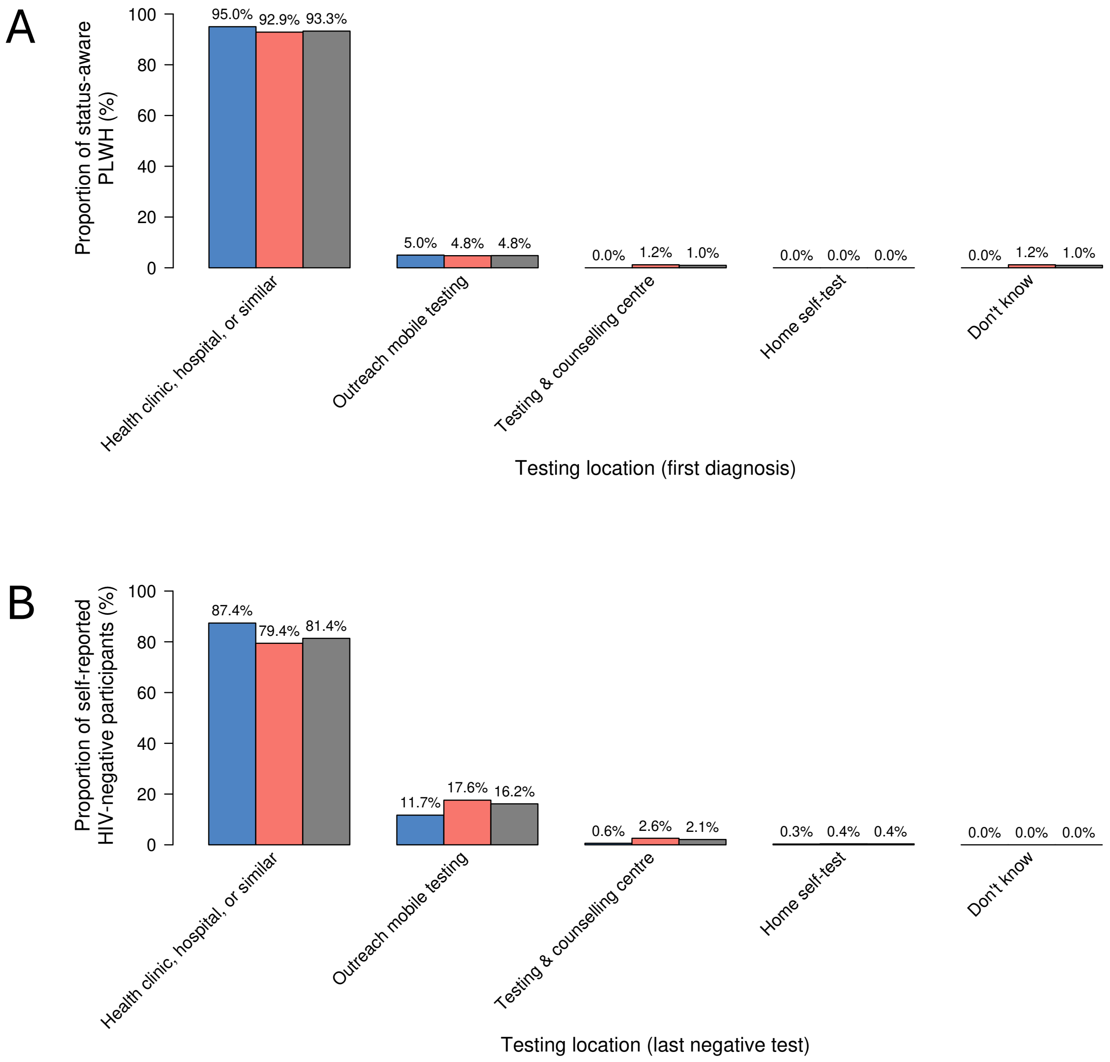

**Figure S3. Testing locations.** First diagnosis (status-aware participants) and last negative HIV test (self-reported HIV-negative participants), stratified by fishing activities (blue – fishing activities; red – no fishing activities; grey – overall). Data are based on participant self-reports from the HIV survey.

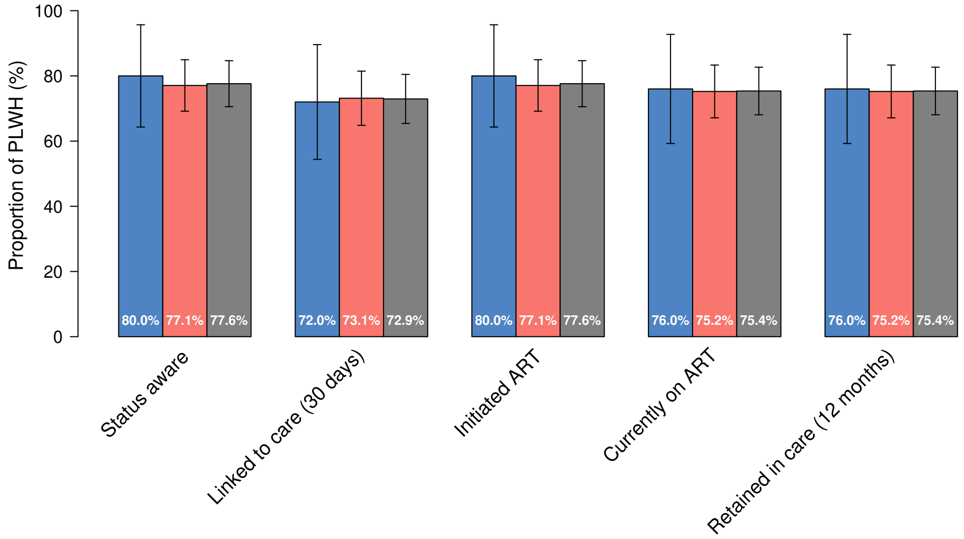

**Figure S4. Expanded diagnosis and care cascade by fishing activities.** Proportion of people living with HIV (PLHIV) at each care cascade stage among all PLHIV (obs.= 134, grey), PLHIV reporting fishing activities (obs.= 25, blue), and PLHIV not reporting fishing activities (obs.= 109, red). Error bars represent Wald 95% confidence intervals. ART = antiretroviral therapy. Data are based on participant self-reports from the HIV survey. One participant (not reporting fishing activities) was missing information on linkage to care and was excluded from the corresponding denominator. Among PLWH who reported previously testing for viral load and knowing the result, viral suppression was reported in 93.44% (57/61) of all PLWH, 100.00% (8/8) of participants reporting fishing activities, 92.45% (49/53) of participants reporting no or other fishing activities.

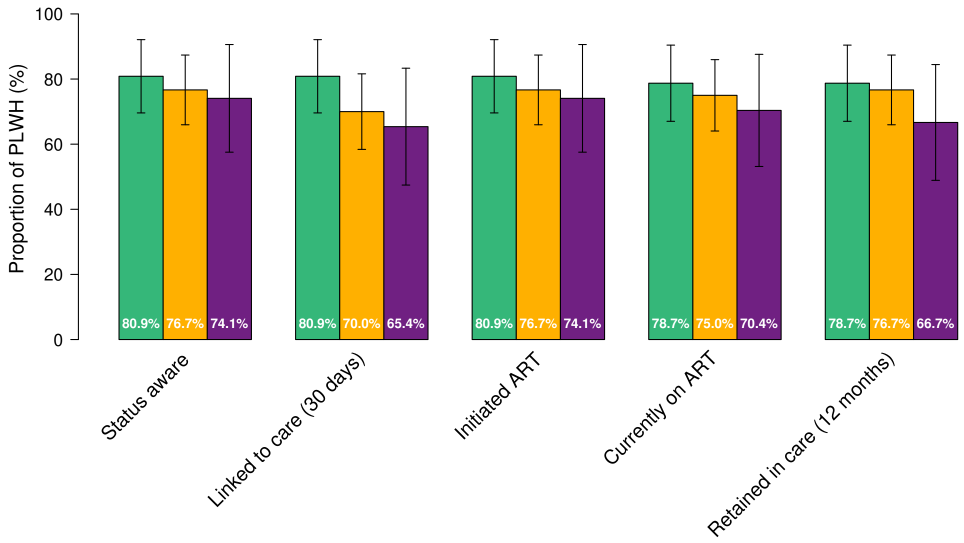

**Figure S5. Expanded diagnosis and care cascade by district.** Proportion of people living with HIV (PLHIV) at each care cascade stage in Buliisa (obs.= 47, green), Mayuge (obs. = 27, purple) and Pakwach (obs.= 60, yellow). Error bars represent Wald 95% confidence intervals. ART = antiretroviral therapy. Data are based on participant self-reports from the HIV survey. One participant (from Mayuge) was missing information on linkage to care and was excluded from the corresponding denominator. Among PLWH who reported previously testing for viral load and knowing the result, viral suppression was reported in 100% (10/10) of participants from Mayuge, 89.29% (25/28) of participants from Buliisa, and 95.65% (22/23) of participants from Pakwach.

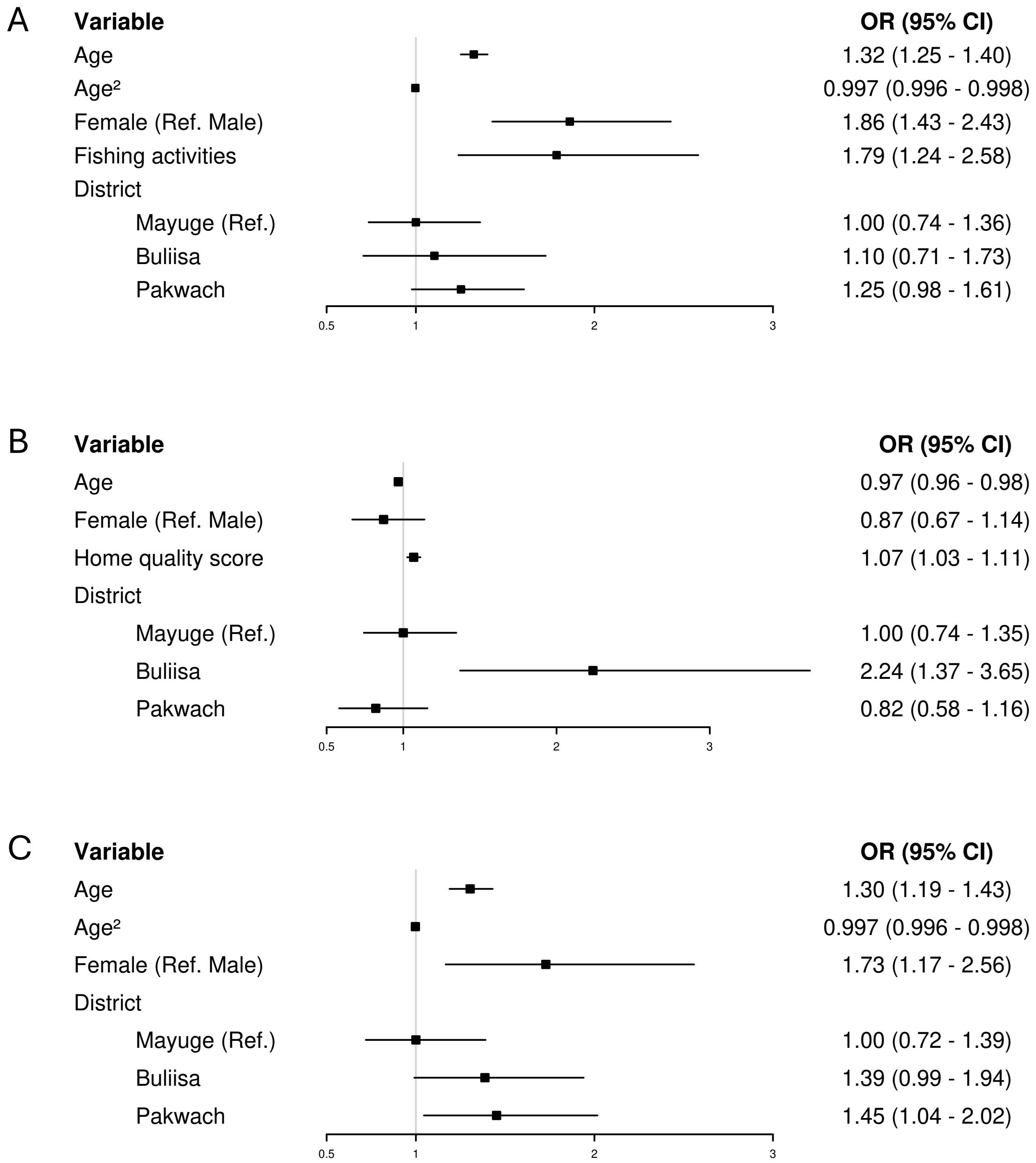

**Figure S6. HIV testing and status models (sensitivity analysis).** (A) Model of ever testing for HIV (obs. = 1935). (B) Model of testing for HIV in the past 12 months among self-reported HIV-negative participants (obs. = 1331). (C) Model of HIV infection (obs. = 1935). Logistic regression models selected by backward stepwise selection based on the lowest Bayesian Information Criterion. Participants who did not know their HIV testing history (ever or in the past 12 months) were included as non-tested. 95% confidence intervals were calculated using village-level clustered standard errors (number of village clusters = 52). Floating absolute risks were calculated for the district variable. ICC = 0.039 (ever testing); 0.23 (testing recency); 0.023 (HIV infection). VIF range: 1.07-25.43 (ever testing); 1.02-1.43 (testing recency); 1.02-34.37 (HIV infection). AUC for stratified ten-fold cross-validation = 0.75 (ever testing); 0.67 (testing recency); 0.65 (HIV infection).
